# preSCRIPT: Large-scale prescription search and annotation engine for pharmacogenomic studies

**DOI:** 10.64898/2026.04.28.26351989

**Authors:** Maria Pieczarka, Paweł Pieńkowski, Paula Konowalska, Sylwia Grubarek, Jacek Hajto, Dzesika Hoinkis, Marcin Piechota, Małgorzata Borczyk, Michał Korostyński

**Affiliations:** Laboratory of Pharmacogenomics, Maj Institute of Pharmacology, Polish Academy of Sciences, Kraków, Poland

**Keywords:** electronic health records, pharmacogenomics, CYP, HLA, GWAS

## Abstract

Pharmacogenetics (PGx) has traditionally focused on a small number of high-impact variants affecting drug response due to the fact that PGx studies are labor-intensive and therefore low-throughput. Population biobanks linked to electronic health records (EHRs), including the UK Biobank (UKB) with prescription data for ∼230,000 individuals offer opportunities to scale PGx research. This, however, comes with a challenge as EHRs do not provide direct treatment response outcomes. One way to overcome this is to draw indirect drug response phenotypes from prescription records. Here, we propose preSCRIPT, a framework to filter and annotate raw prescriptions from the UKB to derive phenotypes for analyses which includes an algorithm to distinguish short prescription gaps from true dose changes. As a proof of concept, we applied preSCRIPT to warfarin, paracetamol, codeine, amitriptyline, simvastatin, aspirin, and amlodipine and derived therapy length and median daily doses. We tested associations for those seven drugs and two phenotypes across SNPs, cytochrome P450 (CYP) genes, and HLA alleles. We replicated known associations such as *CYP2D6* variants with amitriptyline therapy length and dose, *CYP2C9*/*CYP4F2*/*CYP2C19* with warfarin dose, and *CYP2D6* with codeine dose. For drugs without formal PGx guidelines, we identified an association between CYP2D6 activity and aspirin therapy length and several SNPs, including rs62471929 (*CYP3A5*), a variant for amlodipine dose, replicated in an independent hold-out set. Overall, our study shows that preSCRIPT can recover established PGx associations, prioritize exploratory novel candidate loci, and may serve as a tool for large-scale pharmacogenomics.

## Introduction

Traditionally pharmacogenetics (PGx) used to rely on small-scale candidate gene studies. The clinically actionable evidence available today, although successful, mostly rests on a handful of high-impact variants studied gene by gene [1,2]. Implementation is guided by the Clinical Pharmacogenetics Implementation Consortium (CPIC) and curated through resources such as ClinPGx (formerly PharmGKB) [3,4]. In recent years population biobanks linked to longitudinal primary-care records have offered a potential breakthrough in PGx and their use has been suggested as a new frontier for the discipline [5]. UK Biobank (UKB) [6,7], the Estonian Biobank [8], FinnGen [9] and All of US [10] collectively make pharmacogenetic questions tractable on hundreds of thousands of participants. In the UKB alone, 23.7% of participants have already been prescribed at least one drug for which CPIC predicts a non-typical response, and 9.8% a drug for which CPIC recommends an alternative entirely [11]. We have shown that adverse outcomes of antidepressants in the UKB are partially predictable with known PGx variants in *CYP2C19* [12].

The methodology to extract high-quality drug response phenotypes from primary care records, where the only direct read-out of a clinical decision is the prescription, is a recognised barrier to wider EHR-based pharmacology [13]. Towards this, several groups have built tools to extract prescription information from UKB primary-care data. Darke et al. [14] curated a longitudinal research resource that classifies prescriptions by therapeutic group using BNF and Read v2 prefix codes. The T-Rx toolbox of Lo et al. [15] processed around three million antidepressant and antipsychotic prescriptions through a regex-based pipeline anchored on user-supplied substance names. McInnes and Altman [16] used the PharmGKB nomenclature to extract 200 drugs from ∼200,000 UKB participants, while Sadler et al. [17] used UKB and All of Us prescription data for cardiometabolic pharmacogenetics, with a manually curated dictionary of BNF and Read v2 codes.

Condition-specific pipelines exist for treatment-resistant depression [18] and antidepressant change patterns [19]. Although each one of these tools has multiple strong points and use cases, none of them runs from raw UKB primary-care records directly through to ready-to-analyse drug-response phenotypes for any user-defined drugs and is built to run natively on the UKB RAP, the DNAnexus-hosted environment that approved researchers are now required to use.

To validate our prescription extraction framework we analysed seven common drugs and two prescription-derived variables - the dose at which a patient is maintained and the duration over which a treatment is sustained. From the work of others, we know that these phenotypes may carry pharmacogenetic information, even though neither directly measures drug exposure or response. Bijl et al. [20] showed that *CYP2D6*\*4 carriers were prescribed higher doses of antidepressants and switched or discontinued more often than non-carriers, and Lavertu et al. [21] demonstrated that *LPA* and *APOE* genotypes influenced statin selection in UKB. Both findings indicate that prescribing decisions reflect underlying genetics. *SLCO1B1*5* carriers experience elevated rates of atorvastatin discontinuation and adverse muscle symptoms [22–24]. In UKB specifically, Wong et al. [25] recovered an association between CYP2C19 activity score and escitalopram treatment length, a result subsequently extended to antidepressant side effects in the Estonian Biobank [26]. Both pharmacogenomic dose effects and longitudinal medication-use patterns have been shown to have underlying genetics in the Estonian Biobank and FinnGen, including loci unrelated to the therapeutic targets themselves [27,28]. Together, these studies establish that prescription-derived phenotypes carry a recoverable pharmacogenetic signal.

preSCRIPT which we present here is an end-to-end pipeline that converts raw UKB primary-care prescription records into two analysis-ready drug-response phenotypes - median daily dose and longest uninterrupted therapy duration - for a user-defined drug list, and runs natively on the UKB RAP. The current release covers 356 cardiovascular and central-nervous-system (CNS) drugs, but is extendable to any drugs present in the BNF [29]. As proof of concept, we apply preSCRIPT to seven drugs (warfarin, paracetamol, codeine, amitriptyline, simvastatin, aspirin and amlodipine) and test their phenotypes against imputed CYP activity scores, classical HLA alleles, and ∼1.5 million WGS-derived SNPs [30]. We recover well-characterised gene-drug pairs and identify a number of candidate novel signals. We close by comparing the behaviour of dose- and duration-based phenotypes in that hold-out, with implications for how prescription-derived phenotypes can be interpreted in future biobank-scale pharmacogenetic work.

## Methods

### Study population

The UKB is a cohort of 501,946 adults, of whom 44.2% have linked primary care records. We restricted the study cohort to unrelated individuals of self-reported white British ethnicity (field 21000) with genetically confirmed European ancestry (field 22006). Participants flagged for abnormal heterozygosity or elevated genotype missingness (field 22027) or with evidence of sex chromosome aneuploidy (field 22019) were excluded. Participants were pruned to remove closely related individuals at a kinship coefficient > 0.0884 (approximately second-degree relatedness). Kinship was estimated using the KING-robust method; for each pair, the individual with at least one prescription record and, where available, *CYP2D6* genotype data was preferentially retained. The final cohort was randomly partitioned into discovery (80%) and hold-out (20%) subsets before analysis.

#### Genetic data

Three categories of genetic data were processed: (i) pharmacogene activity scores, (ii) HLA allele genotypes, and (iii) genome-wide SNP genotypes. All derived variables were exported in VCF format and converted to PLINK 2 binary format [31] for association testing.

### Pharmacogenes (CYP)

Star-allele diplotypes for *CYP2B6, CYP2C9, CYP2C19, CYP3A5*, and *CYP4F2* were called from UKB whole-exome sequencing pVCFs using PGxPOP [32]. *CYP2D6* diplotypes were obtained from DRAGEN whole-genome *CYP2D6* genotype calls (500k release), reflecting improved calling at the complex *CYP2D6* locus. Star-allele functional annotations were obtained from PharmVar v6.2.14 (June 2025) [33], and each allele was assigned a function category. Per-allele Activity Scores (AS) were then assigned according to PharmVar definitions: normal function = 1.0, decreased = 0.5, no function = 0.0, increased = 2.0; unknown or uncertain alleles were set to normal function. Diplotype AS was computed as the sum of individual allele scores; copy-number increases were handled by multiplying allele scores, while multiple possible diplotype calls were averaged. We manually addressed two edge cases: *CYP2B6*16* was assigned AS = 0.0, and deprecated *CYP3A5* allele labels (*2, *4, *5) returned were recoded as *3 allele. For association testing, each gene’s activity was encoded as a dosage variable DS = AS/2, capped at 2.0, and stored in VCF dosage fields.

### HLA

HLA genotyping data from microarray analysis (362 variants, UKB data-field 22182) were standardized to IPD-IMGT/HLA nomenclature (v3.60.0) [34]. For each HLA type allele values were coded as diploid hard calls (0, 1, or 2) representing homozygous absence, heterozygous presence, and homozygous presence, respectively. Each HLA allele was treated as a biallelic variant on chromosome 6 and exported in VCF format. Analyses were restricted to HLA alleles with a carrier count ≥ 3 for each drug.

### SNPs

Variants from whole genome sequencing mapped to GRCh38 (UKB data-field 24310) were restricted to biallelic SNPs after excluding multi-allelic sites and variants overlapping RepeatMasker-annotated repetitive regions. Variants were retained if call rate ≥ 95%, minor allele frequency > 0.5%, and Hardy-Weinberg equilibrium (HWE) p > 0.01 and all gnomAD [35] QC filters. This resulted in a set of 1,502,697 relatively common and high-quality SNPs. Hemizygous variants on chromosome X were recoded into homozygotes.

#### Phenotypic data

### Prescription processing

Raw prescription records were obtained from the UKB Primary Care Linked Data. Each record contains an issue date, a drug identifier (Read v2, BNF, or dm+d code), and, for 87.0% of entries, a free-text drug name with quantity. In total, 56.2 million records for 221,890 participants were available (data up to September 2017). Records originated from four GP system suppliers: England TPP (69.0%), Wales EMIS/Vision via SAIL (12.9%), England Vision (10.5%), and Scotland EMIS/Vision (7.6%). For preSCRIPT we processed all substances from BNF chapters 02 (Cardiovascular System) and 04 (CNS) defined using NHS Business Services Authority drug sales data for 2023/2024 [36]. After merging entries sharing the same BNF Chemical Substance, 385 drugs were identified, with 356 unique active pharmaceutical ingredients (hereafter, substances).

We used the UKB primary care lookup tables (Resource 592 https://biobank.ndph.ox.ac.uk/showcase/refer.cgi?id=592) where we matched each substance with the BNF Chemical Substance and BNF Product columns. This produced a dictionary with each substance mapped to all available brand names and synonyms. Then we searched all of these names in the term description fields of the UKB lookup tables for all three coding systems (bnf_lkp, read_v2_drugs_lkp, and dmd_lkp sheets accordingly) to identify prescribing codes linked to each substance. Combination products were duplicated for each of their component substances, such that the same code or brand name could be linked to more than one substance where appropriate. Prescriptions were then matched by exact code matching which found 8.4 million records. For unmatched prescriptions we searched the free-text against substance, synonyms and brand names (19.9 million additional matches), retaining 28.4 million prescriptions for CNS and cardiovascular drugs (50.5% of the raw dataset). Prescriptions that matched two or more substances were termed “combination products” and these were split into separate substance-level prescriptions (5.3% of retained entries).

Dose information was extracted from free-text fields (e.g., “40 mg tablets”) using regular expressions and standardized to micrograms. Quantity fields were parsed to extract the total quantity supplied (e.g., number of tablets) per prescription. Records with missing doses or quantities were imputed with the most common value for the given substance, if available. Prescriptions with missing dose and quantity were removed, leaving 24.0 million records (84.5%) for 175,268 participants. This group was then overlapped with a genetic sample QC (see the Study Population section).

### Drug response phenotypes

First, the daily dose for each prescription was estimated by dividing the total amount supplied (dose × quantity) by the interval to the next prescription (days). If another prescription occurred within 14 days, it was combined with the previous one into a single prescription. We then joined adjacent prescriptions into dosing periods. A new dosing period was initiated when the median estimated daily dose changed by more than two standard deviations within a sliding window of five consecutive prescriptions or when the gap between prescriptions was greater than 60 days + quantity (e.g. number of tablets) from the last prescription. For each dosing period, the final daily dose was then calculated by dividing the total supplied amount by the length of the period. Dosing periods without breaks over 60 days were grouped into continent-based treatment courses, which we call therapies. For each substance, two phenotypes were derived: (i) median daily dose across all dosing periods and (ii) longest therapy duration.

For quality control, the longest therapy durations were capped at the mean plus eight standard deviations. Daily doses below one-quarter of the 1st percentile or above four times the 99th percentile were removed as probable data entry errors with remaining values exceeding the mean plus eight standard deviations were also excluded as outliers. All phenotypes were log-transformed using ln(x + 1) to reduce right skewness.

### Covariates

All association models included year of birth (field 34, converted to a continuous timestamp), sex (field 31), the first 16 genetic principal components (field 22009) [37], and a composite education score derived by combining country-specific indices from England (field 26414), Wales (field 26421), and Scotland (field 26431) after percentile normalization within each country, to account for potential geographic and socio-environmental confounding [38]. The primary prescription data provider, defined as the supplier contributing the largest share of each participant’s records, was additionally included as a covariate to account for source-specific prescribing and completeness differences.

### Statistical analysis

Association testing was performed using the generalized linear model framework in PLINK 2.0 with log-transformed phenotypes as outcomes and covariates as described above. Analyses for each drug were conducted within subcohorts of discovery-set participants with at least one prescription for the respective substance. CYP and HLA associations were corrected for multiple testing using the Benjamini-Hochberg procedure (FDR < 0.1), applied independently per phenotype. SNP-based GWAS results were evaluated at genome-wide significance (p < 5 × 10^−8^) and a suggestive threshold (p < 1 × 10^−6^). For SNPs, LD clumping was performed with PLINK 2.0: index variants were defined at p ≤ 1 × 10^−4^, and variants within ± 1,000 kb with r^2^ ≥ 0.2 and p ≤ 1 × 10^−2^ were assigned to the same clump. SNPs were mapped to genes using the Ensembl Variant Effect Predictor (VEP) [39]; variants that could not be annotated this way were assigned to the nearest protein-coding gene based on GRCh38 genomic coordinates retrieved from Ensembl BioMart [40]. All analyses were repeated in the hold-out set. An association was considered replicated if p < 0.05 (nominal) and the direction of effect was consistent.

### Overlap with existing ClinPGx guidelines

Results of associations with analysed CYP and HLA alleles, were cross-referenced with ClinPGx [4] clinical and variant annotations. For CYP genes we mapped each gene-drug pair to ClinPGx annotation with the highest level of evidence. For HLA loci, we matched each HLA type-drug pair where possible (19 out of the 23 associations). If an exact match was not available, we matched on the first two digits (e.g. *HLA-A*:03), and where this was unavailable, we assigned the annotation from the gene-level allele with the lowest p-value from our data.

## Results

### preSCRIPT: Robust prescription capture and annotation from UK Biobank primary care records

The UK Biobank primary care prescription database has 56.2 million records, each annotated with one or two drug identifiers from three coding systems (BNF, Read v2 and dm+d) as well as with three free text fields. Our extensive analysis of the prescription database showed that relying either on coded identifiers or on text fields alone is insufficient for robust prescription retrieval. In the BNF-coded subset (43 million prescriptions), 90.2% of records carry BNF subparagraph codes, which indicate broad drug categories rather than specific products (e.g. Diuretics with Potassium) and only 5.0% of prescriptions have full-resolution BNF codes. Moreover, in the dominant provider TPP (69.0% of all prescriptions), BNF codes are not stored in standard format but are systematically distorted; for example, a code recorded as “04.08.01.03.00” actually represents the BNF subparagraph “0408013”. In dm+d, 76.7% identifiers cannot be resolved using UKB-provided code dictionaries (Resource 592 https://biobank.ndph.ox.ac.uk/showcase/refer.cgi?id=592) because they are in SNOMED CT coding rather than dm+d.

To tackle these and other data irregularities we developed preSCRIPT, a framework for extracting high-quality prescription-derived phenotypes from the UK Biobank primary care records (Fig. 1A, github.com/ippas/prescript). preSCRIPT includes both improved code-matching as well as extensive matching of the free-text drug name field, which is available for 87.0% of prescriptions, as an additional source of information for drug identification. At the center of preSCRIPT are large dictionaries that not only map substances to codes (229.1 per substance on average), but also include brand names and synonyms (7.7 per substance on average).

**Figure 1.**
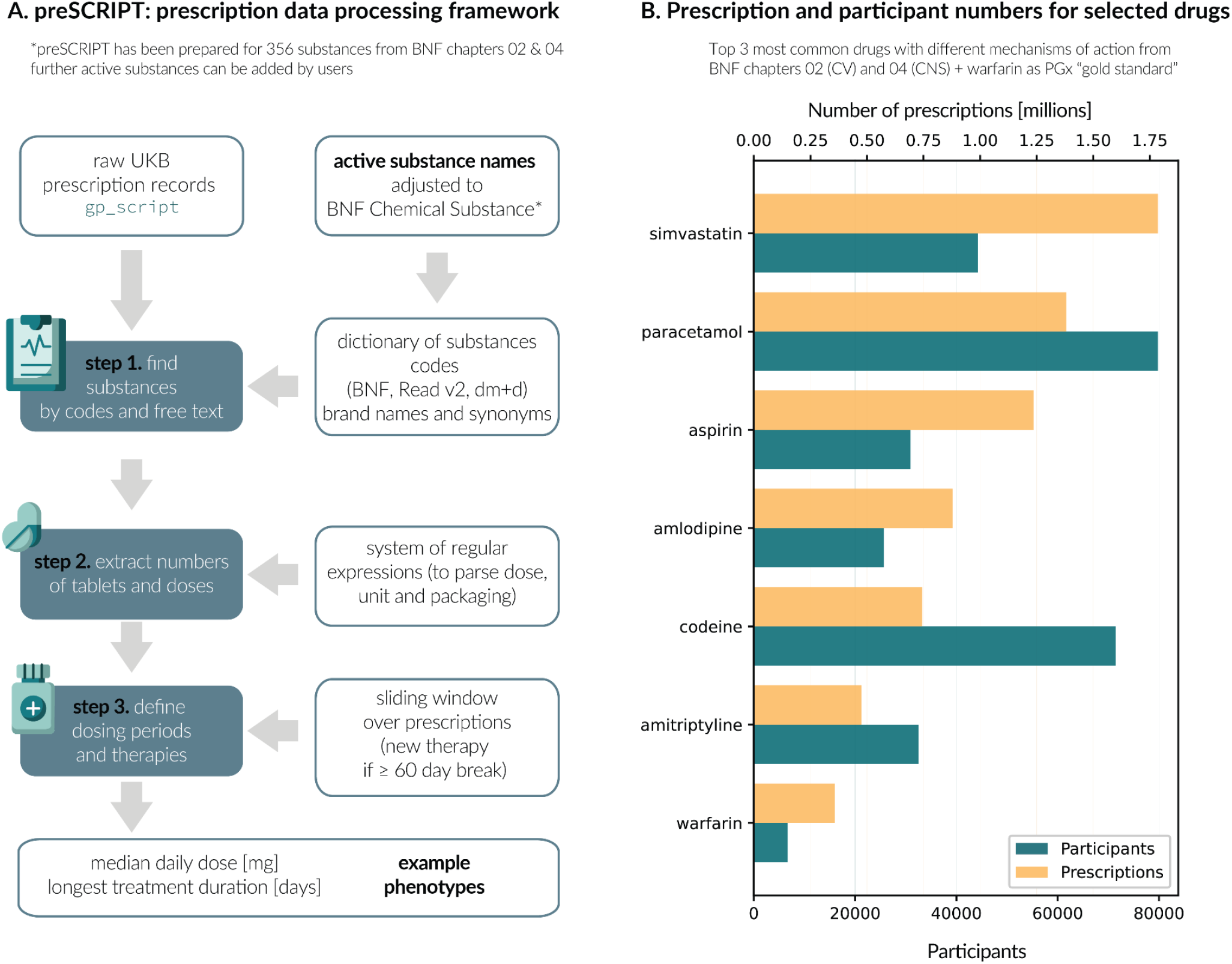
Overview of the prescription data processing framework PRESCRIPT. (A) preSCRIPT pipeline for processing raw UK Biobank prescription records into dosing periods, therapy episodes, and other dose-derived phenotypes. Within it active substances are mapped using codes and free-text search against a modified BN-derived dictionary, doses and tablet counts are extracted using regular expressions and treatment episodes are delineated by applying a sliding window over prescription dates. (B) To validate the utility of preSCRIPT for large-scale PGx studies, we perform genetic association analyses for seven commonly prescribed drugs: paracetamol, codeine, simvastatin, amitriptyline, aspirin, amlodipine and warfarin. The bar chart shows the number of extracted prescriptions (yellow bars, upper x-axis) and the number of participants contributing to each drug-specific group (green bars, lower x-axis).

We ran preSCRIPT for 356 substances, for which code matching alone extracted 8.4 million of 56.2 million UK Biobank prescription records (14.9%), whereas text-based matching recovered a further 19.9 million prescriptions. This increased the total to 28.4 million matched prescriptions (50.5% of all 56.2 million records) for 221,270 participants. Among extracted prescriptions, 1.5 million (5.3%) were combination products containing two or more substances, which we mapped to each constituent substance to avoid losing records.

To validate the framework in association studies, we selected drugs from BNF chapters 02 and 04 with the largest numbers of treated UKB participants, keeping one drug per class to maximize diversity of mechanisms of action. This resulted in a list of three cardiovascular drugs (simvastatin, aspirin and amlodipine) and three neurological drugs (paracetamol, codeine and amitriptyline), together with warfarin as a pharmacogenomic positive control. Within the final study cohort (see Study population section in the Methods), we identified 111,239 individuals taking at least one substance under analysis, including 79,806 for paracetamol (1.38M prescriptions), 71,445 for codeine (743.8k), 44,221 for simvastatin (1.79M), 32,529 for amitriptyline (476.1k), 30,947 for aspirin (1.24M), 25,648 for amlodipine (877.9k) and 6,649 for warfarin (357.1k) (Figure 1B).

### Characterization of prescription-derived drug response proxy phenotypes for seven drugs

A central output of preSCRIPT is a table with one annotated prescription per row. From this table multiple phenotypes can be derived, including traits related to treatment length and dosage. Here we focused on two representative phenotypes: median daily dose and longest therapy duration. Others have already hypothesized that medication dosage can capture altered drug metabolism or treatment resistance [41,42] and the longest uninterrupted therapy duration can reflect therapeutic success understood as both drug efficacy and lack of ADRs [22,43]. preSCRIPT reconstructs individual treatment histories using an algorithm tolerant of minor irregularities, such as imperfect adherence or an occasionally missing prescription (Figure 2A). We first grouped prescriptions into continuous dosing periods and then subsequently into uninterrupted treatment episodes (hereafter, therapies). The procedure yielded 2.5 million dosing periods organised into 859 thousand therapies.

**Figure 2.**
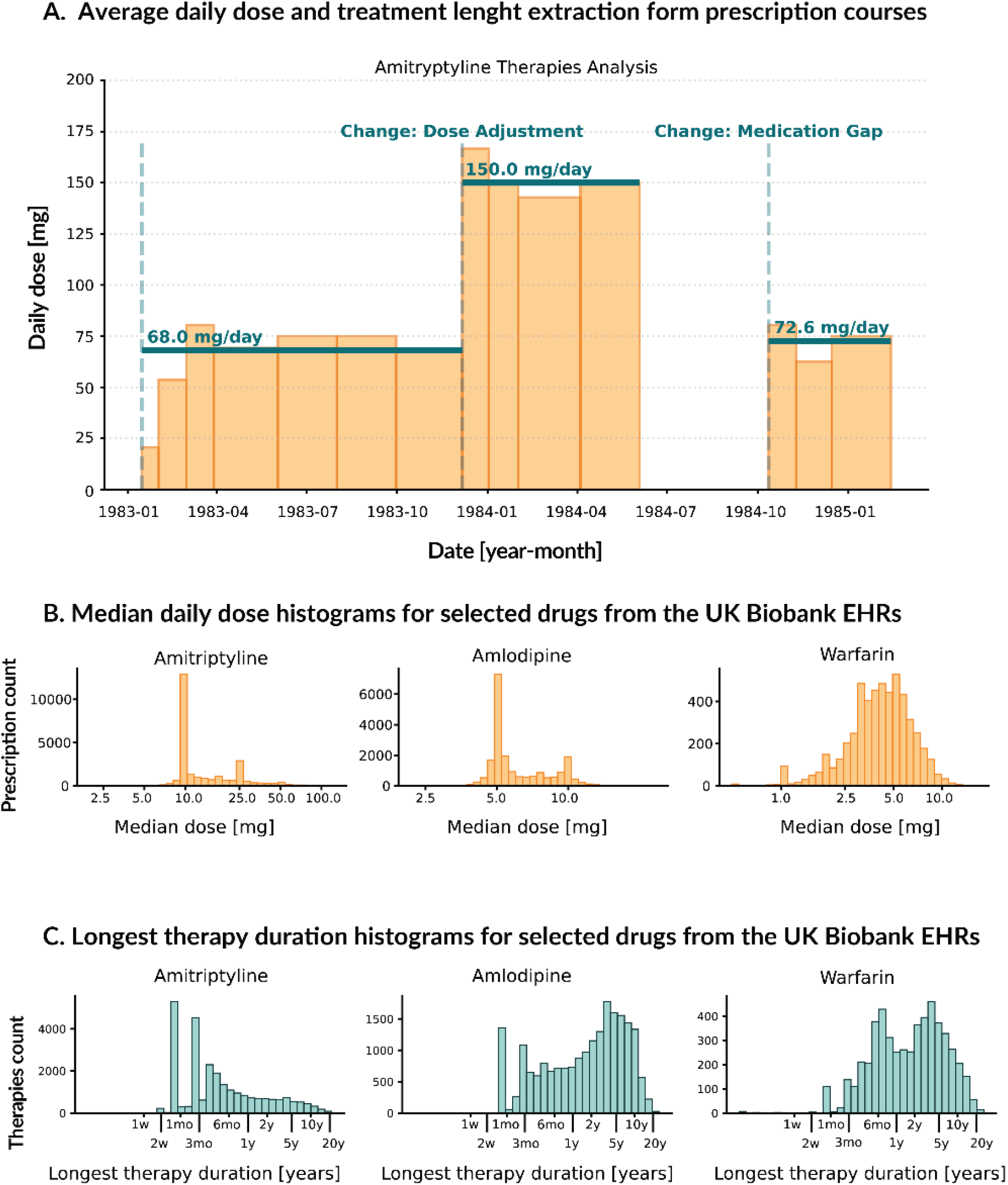
Computation of daily dose and treatment duration phenotypes with preSCRIPT. **A**. Reconstruction of treatment episodes from raw prescription records, illustrated with mock amitriptyline data. Prescriptions were grouped into continuous dosing periods and a new period was initiated when daily dose changed by >2 SD within a sliding window of 5 consecutive prescriptions. Dosing periods were grouped into uninterrupted therapies, with a new therapy defined when the gap between prescriptions exceeded 60 days plus the quantity from the last prescription. **B, C**. Distributions of derived drug-response phenotypes for representative drugs. For each participant, the median daily dose and the longest uninterrupted treatment duration were estimated.

We then analysed the distribution of both doses and therapy durations (Figure 2B and 2C, Supplementary Figure 1). Peaks in the median-dose distributions match commonly prescribed doses: simvastatin 10/20/40 mg [44], aspirin 75 mg (for cardiological purposes) [45], paracetamol 500 mg [46], codeine 8/15/30 mg (sold with paracetamol as co-codamol) [47], amitriptyline 10/25 mg (for neuropathic pain and migraine) [48], and amlodipine 5/10 mg [49]. Longest-therapy distributions, for most drugs, show two short-duration peaks, which are consistent with common pack sizes (30 or 60 tablets). As expected, drugs typically used for long-term treatment, such as simvastatin and amlodipine (Figure 2C, Supplementary Figure 1), have longer right tails of the treatment length than paracetamol or codeine. Warfarin is distinct with approximately normal dose distribution, consistent with individualized dose titration (Figure 2C).

### Prescription-derived phenotypes are associated with known and novel pharmacogenetic variants

As we hypothesise that both treatment length and dose achieved are associated with drug response, we first attempted to replicate existing PGx guidelines for selected drugs. For this we overlapped our results with variant annotations for cytochrome P450 (CYP) and HLA genes from ClinPGx [4] which contains all CPIC recommendations and organises PGx knowledge into evidence levels (from 1A to 3). For our analysis to more closely resemble pharmacogenetic testing results we did not use raw variant data but computed CPIC-style activity scores for six CYP genes (*CYP2B6, CYP2C19, CYP2C9, CYP2D6, CYP3A5, CYP4F2*). Similarly, for the HLA analysis we used the imputed HLA types available through the UK Biobank and tested each type separately, which was 362 HLA alleles in total, including 55 *HLA-A* and 127 *HLA-B*. Out of the seven drugs we analysed, three (amitriptyline, codeine, warfarin) had ClinPGx evidence of level 1 (A or B) for at least one of the analysed genes.

For the six CYP genes we analysed there are 17 known gene-drug associations: 5 at level 1A, 5 at level 3, and 7 annotated at level 4 or with no level. We replicate seven of those at a nominal p-value threshold of 0.05 for either longest therapy duration or optimal dose (Table S6) including four out of five with 1A (Table 1A). These level 1A guidelines that we recovered include the well-established relationship between warfarin dose and CYP2C9 activity (β = 0.66, nominal p-value ≈ 3.4 × 10^−91^) as well as CYP4F2 activity (β = −0.18, nominal p-value = 8.9 × 10^−6^; ClinPGx level 1A). We also recover CPIC guidance for amitriptyline, in our data higher CYP2D6 activity is associated with both higher dose (β = 0.033, nominal p = 1.2 × 10^−3^) and longer treatment duration (β = 0.084, nominal p = 3.1 × 10^−3^), consistent with the recommendation for higher doses in these individuals. For codeine, increased CYP2D6 activity produces greater conversion of codeine to the active metabolite morphine. Consequently, we show that participants with higher CYP2D6 activity are prescribed lower doses of codeine (β = −0.028, nominal p-value = 8.2 × 10^−4^). The only level 1A association that is not visible in our EHR-derived phenotypes is amitriptyline’s association with *CYP2C19* genotypes.

**Table 1.** Associations between CYP scores or HLA alleles and preSCRIPT-derived phenotypes (median daily dose and longest therapy duration). Part A reports all tested ClinPGx level 1A gene-drug pairs. Associations with p-value < 0.05 for any of the two phenotypes are considered replicated. Part B shows lower-level ClinPGx associations that passed the replication threshold. Part C reports novel associations that passed the assumed FDR 10% discovery threshold. Beta values (β) are reported on the ln-transformed phenotype. MAF is only reported for HLA alleles.

| Drug | Gene / variant | MAF | Phenotype | β (ln scale) | Nominal p | Adj. p-value (FDR) | ClinPGx level | Replication |
| --- | --- | --- | --- | --- | --- | --- | --- | --- |
| Table 1A. Replication of ClinPGx level 1A associations, all tested 1A associations listed. |  |  |  |  |  |  |  |  |
| warfarin | CYP2C9 | - | median dose | 0.665 | 3.4 × 10<sup>-91</sup> | 2.0 × 10<sup>-90</sup> | 1A | yes |
| warfarin | CYP2C9 | - | therapy duration | −0.168 | 0.078 | 0.470 | 1A |  |
| warfarin | CYP4F2 | - | median dose | −0.182 | 8.9 × 10<sup>-6</sup> | 2.7 × 10<sup>-5</sup> | 1A | yes |
| warfarin | CYP4F2 | - | therapy duration | 0.074 | 0.527 | 0.941 | 1A |  |
| codeine | CYP2D6 | - | median dose | −0.028 | 8.2 × 10<sup>-4</sup> | 4.9 × 10<sup>-3</sup> | 1A | yes |
| codeine | CYP2D6 | - | therapy duration | −0.021 | 0.139 | 0.520 | 1A |  |
| amitriptyline | CYP2D6 | - | median dose | 0.033 | 1.2 × 10<sup>-3</sup> | 7.3 × 10<sup>-3</sup> | 1A | yes |
| amitriptyline | CYP2D6 | - | therapy duration | 0.084 | 3.1 × 10<sup>-3</sup> | 1.9 × 10<sup>-2</sup> | 1A |  |
| amitriptyline | CYP2C19 | - | median dose | −0.0003 | 0.985 | 0.985 | 1A | no |
| amitriptyline | CYP2C19 | - | therapy duration | 0.037 | 0.313 | 0.618 | 1A |  |
| Table 1B. Replication of ClinPGx level 3 associations, see Tables S6 and S7 for full results. |  |  |  |  |  |  |  |  |
| aspirin | CYP2D6 | - | median dose | 0.0004 | 0.949 | 0.963 | 3 | yes |
| aspirin | CYP2D6 | - | therapy duration | 0.081 | 0.010 | 0.060 | 3 |  |
| warfarin | CYP2C19 | - | median dose | −0.065 | 0.012 | 0.024 | 3 | yes |
| warfarin | CYP2C19 | - | therapy duration | −0.044 | 0.550 | 0.941 | 3 |  |
| simvastatin | CYP2D6 | - | median dose | −0.014 | 0.047 | 0.280 | 3 | yes |
| simvastatin | CYP2D6 | - | therapy duration | 0.002 | 0.924 | 0.924 | 3 |  |
| Table 1C. Candidates for novel associations (FDR < 0.1, not listed in ClinPGx) |  |  |  |  |  |  |  |  |

| Drug | Gene / variant | MAF | Phenotype | $\beta$ (ln scale) | Nominal p | Adj. p-value (FDR) | ClinPGx level | Replication |
| --- | --- | --- | --- | --- | --- | --- | --- | --- |
| amitriptyline | *HLA-DRB3* *01:01 | 0.166 | therapy duration | -0.065 | $2.0 \times 10^{-4}$ | 0.041 | - | novel |
| amitriptyline | *HLA-DRB3* *01:01 | 0.166 | median dose | -0.012 | 0.046 | 0.565 | - |  |
| amitriptyline | *HLA-DQA1* *03:01 | 0.204 | therapy duration | 0.058 | $3.6 \times 10^{-4}$ | 0.041 | - | novel |
| amitriptyline | *HLA-DQA1* *03:01 | 0.204 | median dose | 0.012 | 0.041 | 0.565 | - |  |

Apart from those established associations we also confirm some level 3 evidence from ClinPGx. Namely, individuals with higher CYP2C19 activity are on average prescribed lower warfarin doses (β = −0.065, nominal p = 0.012; ClinPGx level 3, Table S6) and higher CYP2D6 activity is associated with longer aspirin treatment duration (β = 0.081, nominal p = 0.010; ClinPGx level 3, Table S6) and lower simvastatin dosage (β = −0.014, nominal p = 0.047; ClinPGx level 3, Table 1B)

For HLA there are nine unique gene-drug pairs in ClinPGx for our seven drugs. Out of those we replicate three associations for aspirin: with *HLA-DPB1* (β = −0.25, nominal p = 3.7 × 10^−3^; ClinPGx level 3), *HLA-DRB1\**09:01 (β = 0.14, nominal p = 3.0 × 10^−2^) and *HLA-DQB1\**03:03 (β = 0.087, nominal p = 6.5 × 10^−3^), all for the longest therapy duration (Table S7). Overall, the above results show that the UK Biobank EHRs allow for recovery of a substantial portion of known pharmacogenetics associations. What is more, as we analysed all six CYP genes and all HLA types for each of the drugs we were able to identify possible novel associations, which (at an FDR threshold of 0.1) are *HLA-DRB3\**01:01 (β = −0.065, adj. p-value = 0.041) and *HLA-DQA1\**03:01 (β = 0.058, adj. p-value = 0.041) for amitriptyline treatment length (Table 1C).

### Genome-wide association analysis of preSCRIPT-derived therapy lengths and medication doses

As we have confirmed that our phenotypes recover established pharmacogenetic associations, we conducted GWAS across the seven drugs (simvastatin, aspirin, amlodipine, paracetamol, codeine, amitriptyline and warfarin) and our two phenotypes (median daily dose and longest therapy duration) to screen for additional pharmacogenetic signals, both previously reported and novel. After clumping, we identified a total of 12 genome-wide significant loci (p-value ≤ 5 × 10^−8^) and 21 suggestive associations (p-value < 1 × 10^−6^) for dose phenotypes and 2 genome-wide significant and 7 suggestive associations for treatment duration phenotypes (Table S8, for full results see Data Availability section).

Of the 14 genome-wide significant results (Table S8, selected lead variants in Table 2), 11 were associated with warfarin dose. These included the *CYP2C9* locus on chromosome 10 with the lead variant rs9332172 (β = −0.22, p-value = 1.8×10^−77^) and *VKORC1* region on chromosome 16 with rs79030220 ∼2.6 kb upstream of *VKORC1* (β = −0.17, p-value = 1.1 ×10^−18^). Together with the suggestive association of rs8110863 (β = 0.050, p = 6.94 × 10^−8^) which is ∼29 kb downstream of the 3′ end of *CYP4F2* these three peaks repeat the classic warfarin dose associations [50] (Figure 3).

**Table 2.** Selected GWAS associations across the 14 GWAS scans (seven drugs for median daily dose and longest therapy duration). Full results are available in Table S8. β is on the ln-transformed phenotype. MAF is the minor-allele frequency in the analysis cohort. The ClinPGx level column reports the highest ClinPGx evidence level for the closest gene; novel indicates no existing ClinPGx annotation.

| Drug | Phenotype | SNP | Locus (chr:pos) | MAF | $\beta$ | p-value | Gene | ClinPGx level |
| --- | --- | --- | --- | --- | --- | --- | --- | --- |
| warfarin | median dose | rs9332172<br>[A/G] | 10:94,972,031 | 0.196 | -0.22 | $1.8 \times 10^{-77}$ | *CYP2C9*<br>intronic | 1A |
| warfarin | median dose | rs79030220<br>[C/T] | 16:31,088,261 | 0.059 | -0.17 | $1.1 \times 10^{-18}$ | *VKORC1*<br>intronic | 1A |
| warfarin | therapy duration | rs146830815<br>[G/T] | 14:49,413,751 | 0.008 | -0.83 | $1.8 \times 10^{-8}$ | *RPS29*<br>intergenic | novel |
| aspirin | median dose | rs183141088<br>[G/A] | 6:69,138,772 | 0.006 | 0.09 | $2.7 \times 10^{-8}$ | *ADGRB3*<br>intronic | novel |
| aspirin | therapy duration | rs13290723<br>[G/A] | 9:314,464 | 0.236 | 0.09 | $4.2 \times 10^{-8}$ | *DOCK8*<br>intronic | novel |
| warfarin | median dose | rs8110863<br>[T/C] | 19:15,848,595 | 0.407 | 0.05 | $6.9 \times 10^{-8}$ | *CYP4F2*<br>downstream | novel |
| amlodipine | median dose | rs62471929<br>[A/G] | 7:99,676,693 | 0.057 | -0.03 | $2.7 \times 10^{-7}$ | *CYP3A5*<br>intronic | 3 |
| simvastatin | median dose | rs2272642<br>[A/G] | 8:23,436,899 | 0.182 | -0.02 | $3.2 \times 10^{-7}$ | *ENTPD4*<br>intronic | novel |
| amitriptyline | median dose | rs189330<br>[A/T] | 17:15,556,691 | 0.317 | 0.03 | $4.2 \times 10^{-7}$ | *TVP23C*<br>intronic | novel |
| warfarin | median dose | rs1799963<br>[G/A] | 11:46,739,505 | 0.017 | -0.18 | $8.2 \times 10^{-7}$ | *F2*<br>3' UTR | no level |

**Figure 3.**
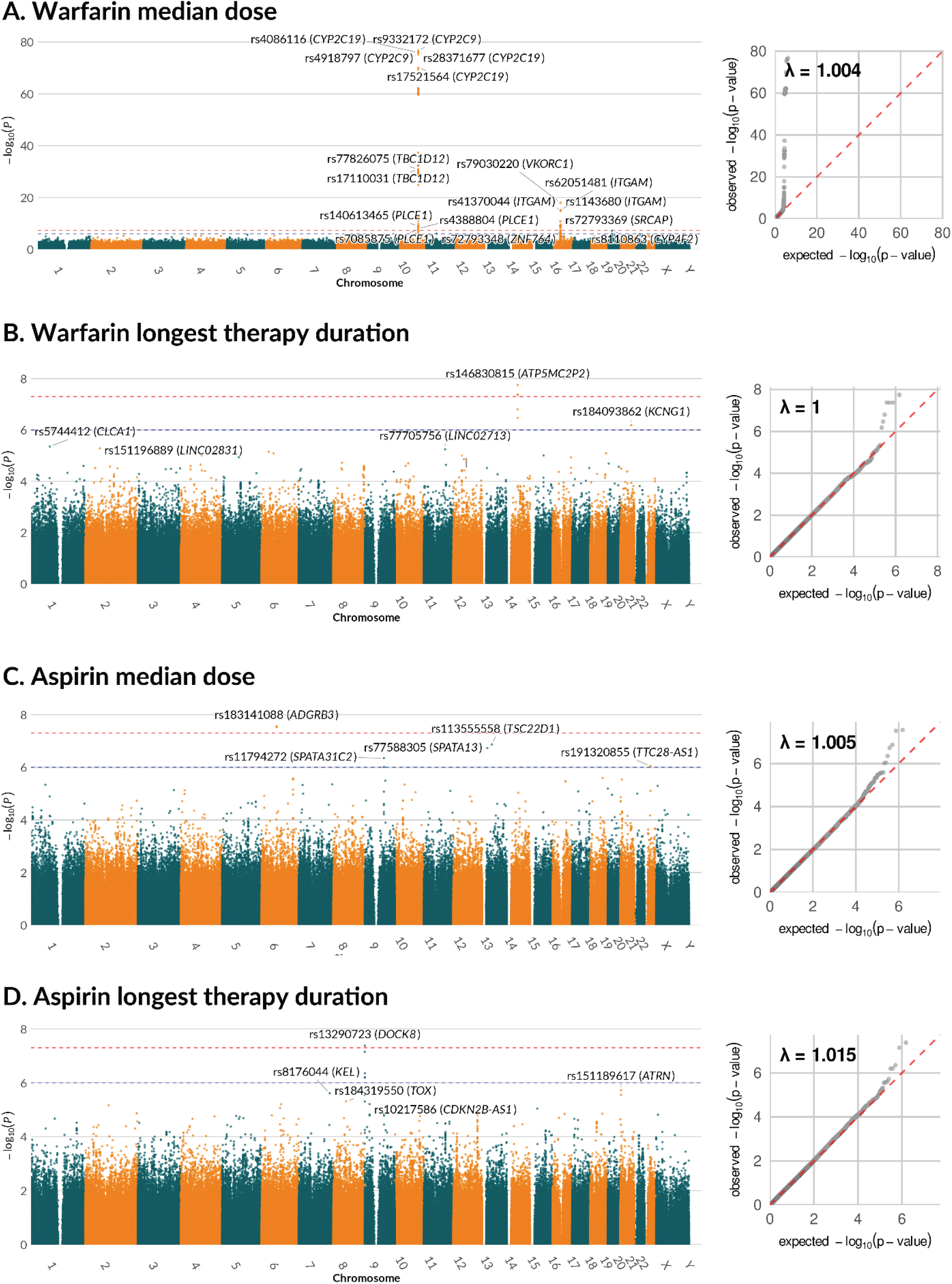
GWAS of median daily dose and longest treatment duration for warfarin and aspirin. Manhattan and quantile-quantile plots for four drug response phenotypes (see Supplementary Figure 2 for the remaining drugs). Dashed lines indicate suggestive (p-value < 1×10-6) and genome-wide significance thresholds (p-value <5 x 10^−8^).

The remaining three genome-wide significant loci were associated with warfarin treatment duration (rs146830815 ∼160kb from *RPS29*, β = −0.83, p = 1.8×10^−8^), aspirin dose (rs183141088 in *ADGRB3*, β = 0.094, p = 2.7×10^−8^), and aspirin treatment duration (rs13290723 in *DOCK8*, β = 0.093, p = 4.2×10^−8^). Examples of suggestive associations (p < 1×10^−6^) include rs1799963 in the 3’UTR of *F2* for warfarin dose (β = −0.18, p = 8.2×10^−7^), a gene in which missense mutations have been long known to alter warfarin requirements [51] as well as rs183575962 within *LRRC27* for simvastatin dose (β = −0.089, p = 2.9×10^−7^) and rs62471929 within *CYP3A5* for amlodipine dose (β = −0.033, p = 2.65×10^−7^).

### Dose phenotypes show a more consistent signal as compared with therapy length in a replication analysis

To assess the stability of the results we repeated the GWAS on a non-overlapping hold-out subset of 36,408 UKB participants. We would like to note that this hold-out set is quite small and thus our replication analysis is underpowered. Therefore, as expected, many associations do not replicate at p-value <0.05. Nevertheless, we report that all dose associations showed a consistent direction of effect in this repeated analysis (Figure 4A, Supplementary Figure 3A). This included associations between warfarin dose and *CYP2C9* (β = 0.67, p-value ∼ 0), *CYP4F2* (β = −0.18, p-value = 9.0×10^−6^), and *CYP2C19* (β = −0.065, p-value = 1.2×10-2), as well as between amitriptyline dose and *CYP2D6* (β = 0.033, p = 1.2×10^−3^), and codeine dose and *CYP2D*6 (β = −0.028, p-value = 8.2×10^−4^). The novel simvastatin dose association rs2272642 (*ENTPD4*), which was suggestive in the discovery (β = −0.021, p-value = 3.2 × 10^−7^) showed a trend (β = −0.014, p-value = 0.081) in the replication analysis. Another finding is rs62471929 (*CYP3A5*), a variant associated with amlodipine dose both in the discovery (β = −0.033, p = 2.7 × 10^−7^) and replication (β = −0.038, p = 3.4 × 10^−3^).

**Figure 4.**
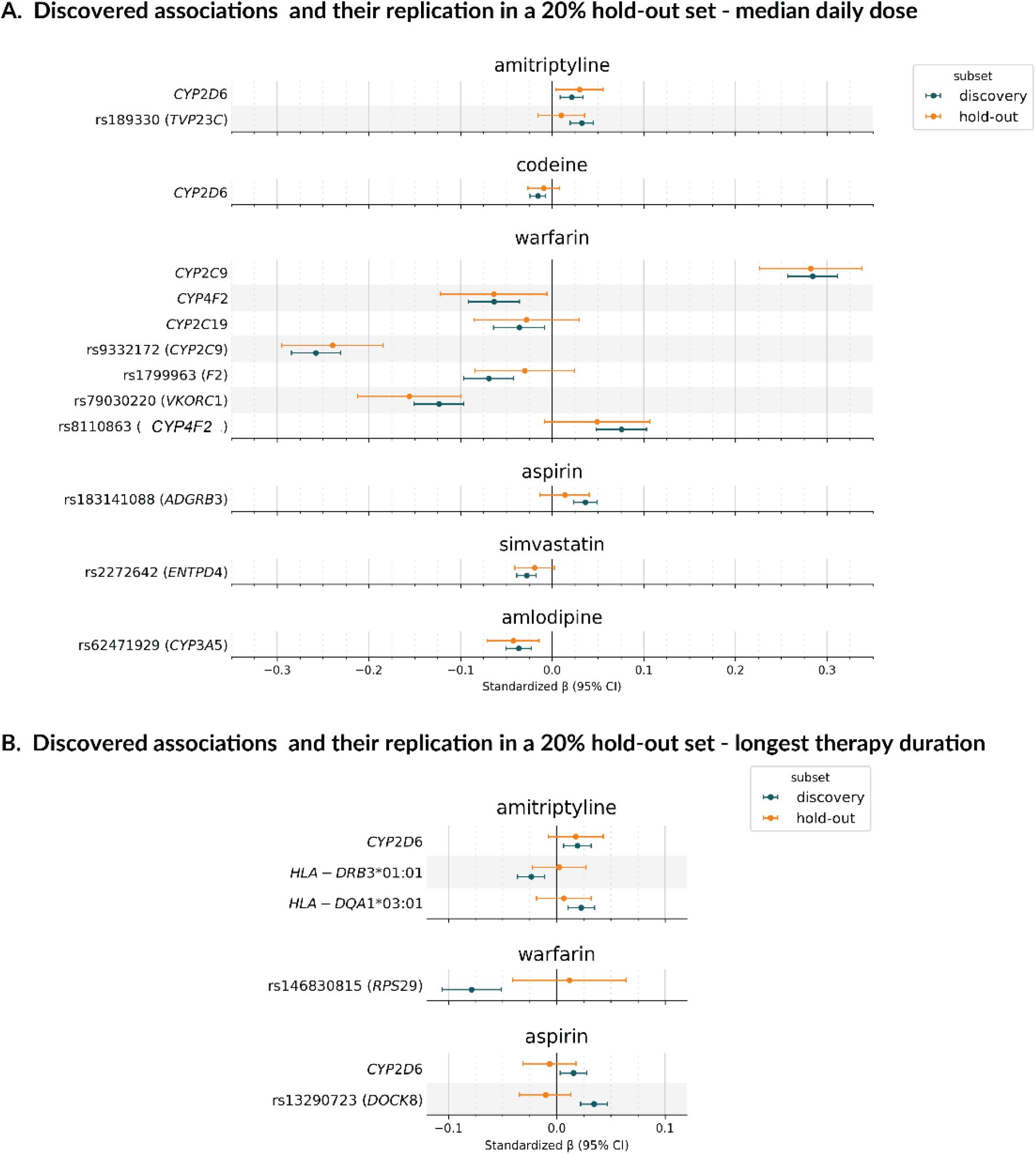
Replication of genetic associations in a hold-out set of 36,408 UK Biobank participants. Each graph shows beta values (ln-transformed) with 95% CI for the discovery set (teal) and the hold-out set (orange). (A) Replication results for median daily dose, (B) longest therapy duration. We tested all associations with FDR <10% from HLA/CYP analysis and p-value < 10 x 10^−6^ from GWAS. Selected associations are plotted here (corresponding to Tables 1 and 2). Full results of the replication are available as Table S9.

From all findings on longest therapy duration, none replicated at p-value < 0.05 and many showed either no effect or effect in the opposite direction (Figure 4B, Supplementary Figure 3B). The closest one to replicate was the known relationship between amitriptyline and CYP2D6 activity (replication β = 0.078, p-value = 0.17). Overall, dose-based associations were the more reproducible of the two phenotypes in our hold-out set, while therapy-length signals lost power or direction.

## Discussion

Here we have presented preSCRIPT, an end-to-end pipeline that converts raw UK Biobank primary-care prescription records into analysis-ready, pharmacogenetically informative phenotypes for any user-defined drug list. As a proof of concept we applied preSCRIPT to seven cardiovascular and CNS drugs and derived two phenotypes per drug: median daily dose and longest uninterrupted therapy duration. Both phenotypes recovered ClinPGx level 1A CYP-drug pairs, supported lower-evidence (level 3) candidate associations, and surfaced a small number of novel signals.

To our knowledge, preSCRIPT is the first publicly available pipeline that converts raw UK Biobank primary-care records into ready-to-analyse phenotypes for an arbitrary, user-supplied drug list and was designed for the UKB RAP. Existing tools each cover only part of their functionalities. Common limitations include the absence of dose extraction, reliance on pre-filtered prescriptions or pre-coded inputs, limited matching between active substance names and prescription codes, and exclusion of combination products [14–18]. preSCRIPT integrates all three UK Biobank coding systems (BNF, Read v2, dm+d) while also matching brand names and synonyms to active substances. It also applies a quantity-aware, dose-aware treatment-episode construction algorithm that distinguishes short prescription gaps from dose changes and it returns ready-to-use phenotypes rather than annotated prescriptions alone. Across our analyses, preSCRIPT-derived phenotypes recovered the principal pharmacogenetic signals expected for the seven test drugs. The genome-wide scan replicated the canonical warfarin loci at *CYP2C9, VKORC1* and *CYP4F2* [50], confirming that prescription-derived dose phenotypes carry sufficient signal for genome-wide replication.

The clinical pharmacogenetics literature for the seven drugs has five level 1A CYP-drug pairs, four of which we replicated at nominal significance in either the dose or the duration phenotype. Apart from Warfarin, Codeine and *CYP2D6* also replicated as expected, with higher CYP2D6 activity associated with lower prescribed doses. This direction is consistent with the CPIC recommendation that clinicians lower codeine doses in CYP2D6 ultra-rapid metabolisers, due to the increased risk of opioid toxicity associated with accelerated conversion of codeine to morphine [52]. For amitriptyline, both median daily dose and treatment length recovered the *CYP2D6* signal: higher activity was associated with higher prescribed dose and longer therapy duration, consistent with the CPIC tricyclic dosing guideline [53]. The only level 1A association that did not reach nominal significance was amitriptyline and *CYP2C19*. The most plausible explanation is the limited resolution of CYP2C19 in our pharmacogene calls. *CYP2C19*17*, an upstream variant associated with rapid or ultra-rapid metabolism [54], lies in a non-coding region and is not reliably captured by whole-exome sequencing.

For less-known associations, we provide population-scale support for the aspirin-*CYP2D6* association previously reported in a 37-case study of low-dose-aspirin-associated small bowel bleeding, with higher CYP2D6 activity associated with longer aspirin therapy duration [55]. The *HLA-DPB1\**03:01 association with aspirin therapy duration, although directionally consistent in our data, did not reach nominal significance in our predominantly European cohort [56,57].

Another finding worth mentioning is the rs62471929 in *CYP3A5*, associated with amlodipine median dose at suggestive significance in the discovery set and replicating in an independent hold-out subset. CYP3A5 genotypes have been linked to altered amlodipine clearance and blood-pressure response in Asian cohorts [58], and the broader effect of CYP3A4 and CYP3A5 variation on amlodipine and other CYP3A substrates has been demonstrated in mixed-population pharmacokinetic studies [59]. The signal in European cohorts has been smaller and less consistent, possibly because the functional *CYP3A5*1* allele is rare in European populations [60]. To our knowledge, our result is the first replicated population-scale support for *CYP3A5* involvement in amlodipine prescribing in a predominantly European cohort.

The novel simvastatin signal at rs2272642 in *ENTPD4* reached suggestive significance in the discovery set but did not replicate at the nominal threshold, although the direction of effect was preserved. *ENTPD4* has no prior pharmacogenetic annotation for statin response and the signal should be regarded as exploratory. The two aspirin signals (rs183141088 in *ADGRB3* for median dose and rs13290723 in *DOCK8* for therapy duration) and the amitriptyline signal at rs189330 in *TVP23C* are similarly novel and require independent replication in larger cohorts before any biological interpretation can be advanced.

Throughout the study, we made a consistent observation. Dose-based associations had more baseline associations and replicated more consistently than duration-based associations in the hold-out subset. One possible explanation is methodological: because we did not exclude short-course prescriptions from the duration phenotype, this readout aggregates several distinct clinical trajectories, including discontinuation due to adverse drug reactions and long-term maintenance therapy reflecting both tolerability and efficacy [20,61]. Different genetic mechanisms drive each of these trajectories, and aggregating them into a single phenotype is expected to attenuate locus-specific signals. Nevertheless, strong pharmacogenetic effects remain recoverable: the amitriptyline-*CYP2D6* therapy-duration signal is consistent with the CPIC-recommended dose-adjustment biology and demonstrates that duration is informative when the underlying effect is sufficiently large.

In summary, preSCRIPT provides a reproducible, RAP-native workflow that converts UK Biobank primary-care prescription records into pharmacogenetically informative phenotypes for any user-defined drug list. The pipeline recovers established CPIC guidelines, contributes population-scale evidence for candidate associations, and identifies novel variants for further studies.

## Data Availability

The UK Biobank has ethical approval from the North West Multi-centre Research Ethics Committee
as a research tissue bank, and all participants provided written informed consent for the use of
their de-identified data in health-related research. These data are available to qualified researchers
upon application (https://www.ukbiobank.ac.uk). The terms of the data access agreement preclude
public sharing of individual-level data.
preSCRIPT code is available at https://github.com/ippas/prescript, while the remaining code used
in this study is available in the repository https://github.com/ippas/ukb-seven-drug-pgx-analyses.
All code is adapted to run on the Research Analysis Platform (RAP), in accordance with its
regulations.
Full GWAS summary statistics are available on the NHGRI-EBI GWAS catalog under the
accession IDs GCST90837327-GCST90837340.

https://github.com/ippas/ukb-seven-drug-pgx-analyses

https://www.ebi.ac.uk/gwas/

## Acknowledgements

The project is funded under the National Recovery and Resilience Plan (KPO) from funds distributed through the Medical Research Agency, Poland, project no.: KPOD.07.07-IW.07-0099/24. Additional support came from the statutory funds of the Maj Institute of Pharmacology PAS.

This research has been conducted using the UK Biobank Resource under Application Number 879030. We are grateful to the UK Biobank and all its voluntary participants. This work used data provided by patients and collected by the NHS as part of their care and support.

This work would not have been possible without the logistical organization and technical expertise provided by Lidia Radwan, Sebastian Plonka, Michal Bozyk and Rafal Kafel.

## Contributions

M.Piecz.: data analysis, methodology, writing - original draft; P.P.: data analysis, methodology, writing - original draft; P.K.: data analysis, methodology; S.G.: data analysis, methodology; J.H.: data analysis, methodology; D.H.: data analysis, methodology; M.Piech.: data analysis, interpretation, conceptualization; M.B.: conceptualization, interpretation, writing - original draft, supervision, writing - review and editing; M.K.: conceptualization, interpretation, supervision, writing - review and editing.

## Ethics approval

The UK Biobank has obtained approval from the North West Multi-center Research Ethics Committee as a Research Tissue Bank approval (references: 11/NW/0382, 16/NW/0274, and 21/NW/0157). This approval covers all researchers using the UK Biobank resource under an approved application; therefore, separate ethical approval was not required for this study. All participants provided written informed consent to participate in the study.

## Data availability

The UK Biobank has ethical approval from the North West Multi-centre Research Ethics Committee as a research tissue bank, and all participants provided written informed consent for the use of their de-identified data in health-related research. These data are available to qualified researchers upon application (https://www.ukbiobank.ac.uk). The terms of the data access agreement preclude public sharing of individual-level data.

preSCRIPT code is available at https://github.com/ippas/prescript, while the remaining code used in this study is available in the repository https://github.com/ippas/ukb-seven-drug-pgx-analyses. All code is adapted to run on the Research Analysis Platform (RAP), in accordance with its regulations.

Full GWAS summary statistics are available on the NHGRI-EBI GWAS catalog under the accession IDs GCST90837327-GCST90837340.

## Abbreviations

ADR: adverse drug reaction
AS: activity score
BNF: British National Formulary
CPIC: Clinical Implementation Pharmacogenetic Consortium
DS: genetic dosage
EHR: electronic health records
HWE: Hardy-Weinberg equilibrium
PGx: pharmacogenomics
RAP: Research Analysis Platform
UKB: UK Biobank

