## Supplementary material for "preSCRIPT: Large-scale prescription search and annotation engine for pharmacogenomic studies"

**Running head:** preSCRIPT: Prescription search and annotation for pharmacogenomics

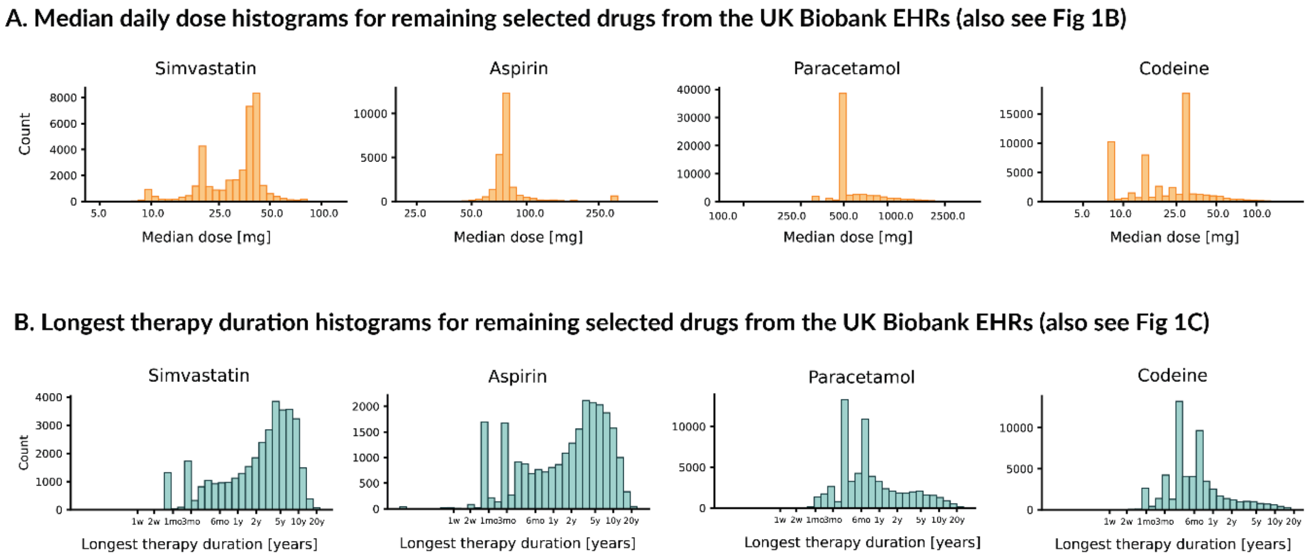


**Supplementary Figure 1. Daily dose and longest therapy duration for four additional selected drugs in the UK Biobank.** Distributions of median daily dose and longest treatment duration (accompanies Figure 2). Phe

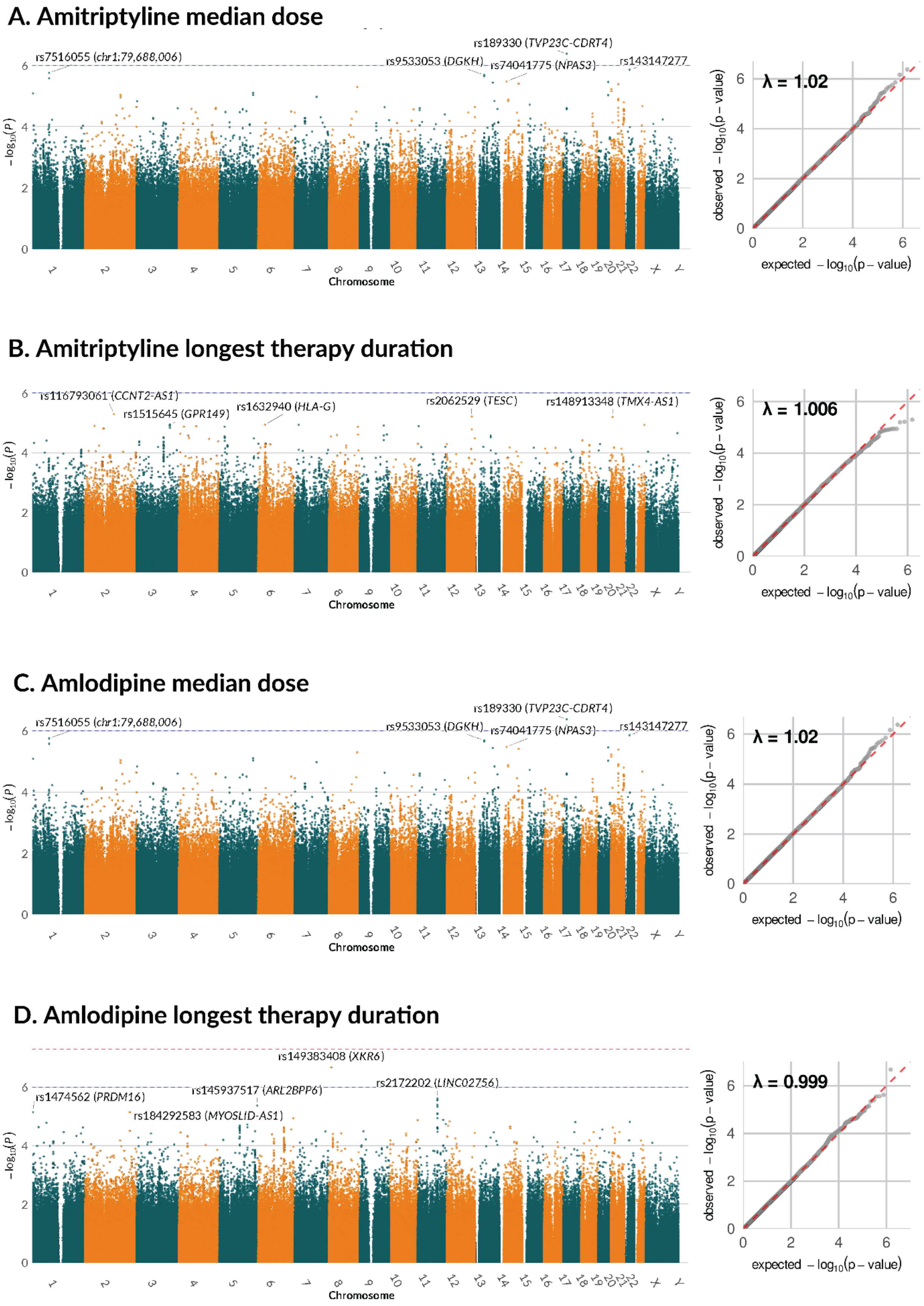


notypes were log-transformed (ln[x + 1]) prior to analysis.

### E. Simvastatin median dose

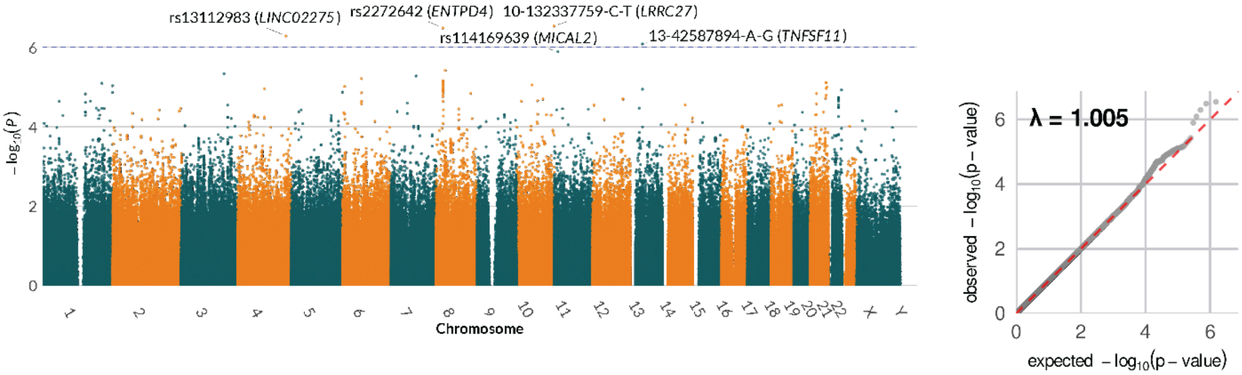


### F. Simvastatin longest therapy duration

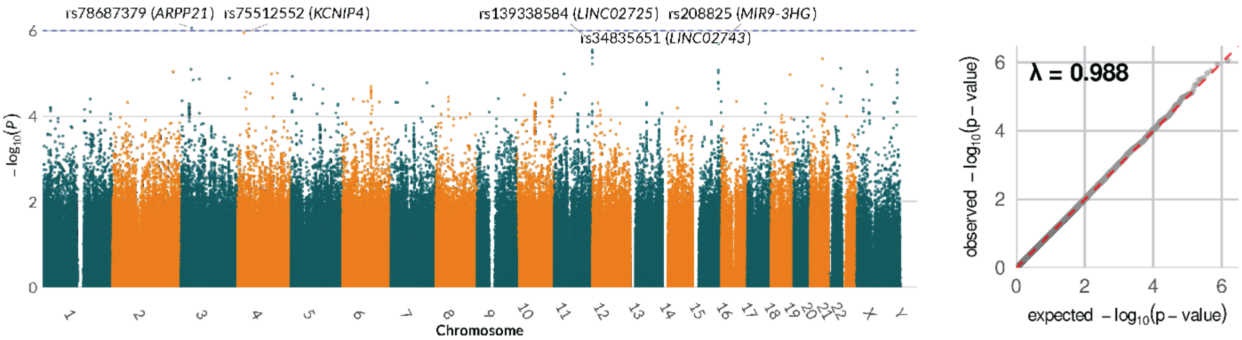


### G. Paracetamol median dose

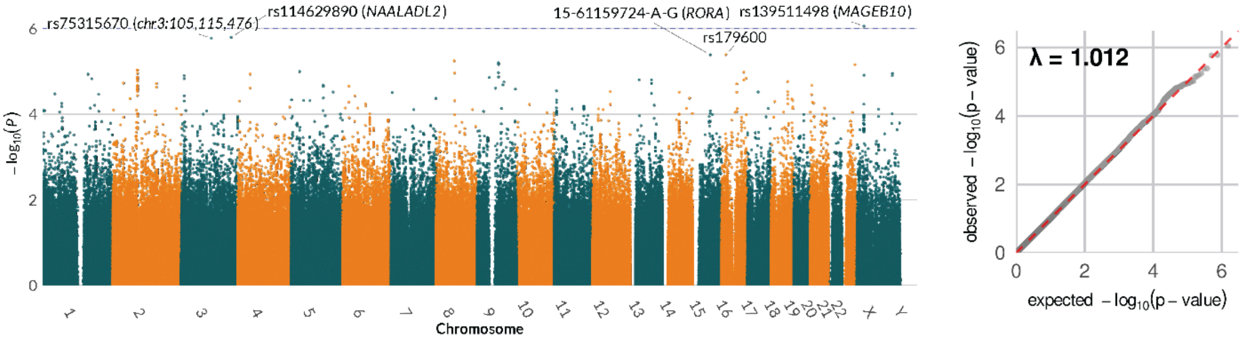


### H. Paracetamol longest therapy duration

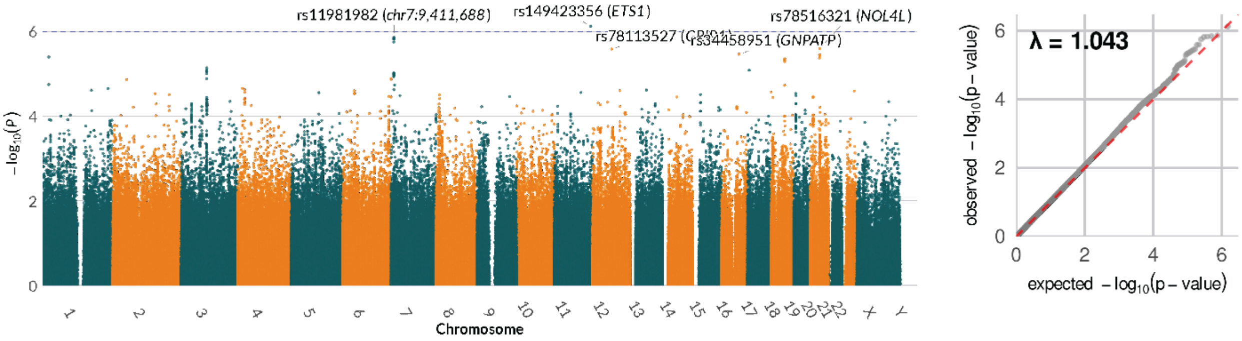

### I. Codeine median dose

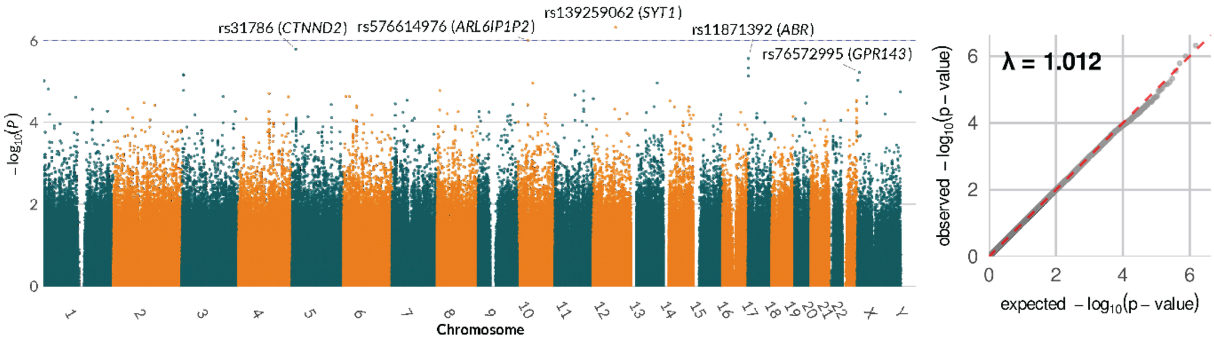


### J. Codeine longest therapy duration

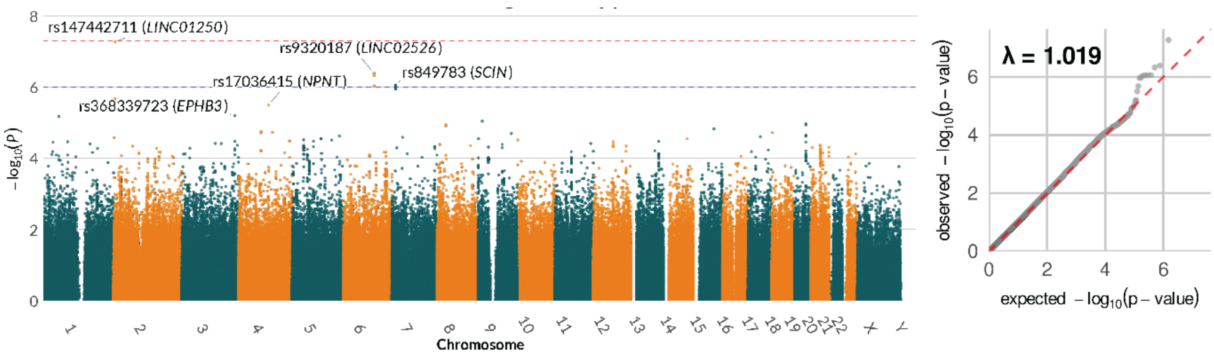


**Supplementary Figure 2. GWAS of median daily dose and longest treatment length (accompanies Figure 3).** Manhattan and quantile-quantile plots for four drug response phenotypes. Dashed lines indicate the suggestive (p-value < $1\times10^{-6}$) and genome-wide significance thresholds (p-value <$5 \times 10^{-8}$).

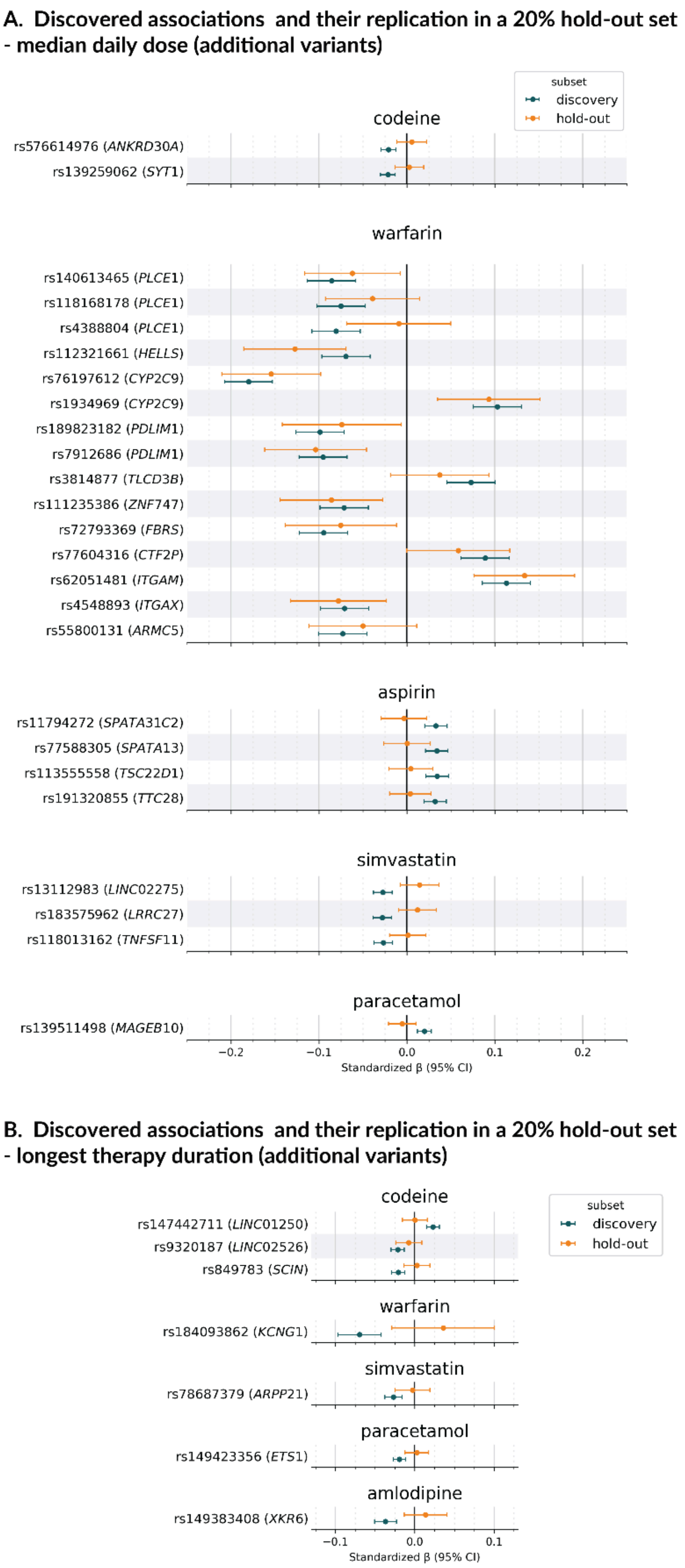


**Supplementary Figure 3. Replication of found genetic associations in an independent hold-out set of 36,408 UK Biobank participants (accompanies Figure 4).** Each panel shows beta values (ln-transformed) with 95% CI for the discovery set (teal) and the hold-out set (orange). We tested all the associations reaching FDR <10% in the HLA/CYP analysis and p-value < 10 x $10^{-6}$ in the GWAS. One representative association per chromosome is shown (corresponding to Tables 1 and 2, see Supplementary Figure 2 for more associations). Full replication results are provided in Table S9.
